# Increased CHIP Prevalence Amongst People Living with HIV

**DOI:** 10.1101/2020.11.06.20225607

**Authors:** Alexander G. Bick, Konstantin Popadin, Christian W. Thorball, Md Mesbah Uddin, Markella Zanni, Bing Yu, Matthias Cavassini, Andri Rauch, Philip Tarr, Patrick Schmid, Enos Bernasconi, Huldrych F. Günthard, Peter Libby, Eric Boerwinkle, Paul J. McLaren, Christie M. Ballantyne, Steven Grinspoon, Pradeep Natarajan, Jacques Fellay, the Swiss HIV Cohort Study

**Author notes:** Correspondence to Dr. Pradeep Natarajan or Dr. Jacques Fellay, Pradeep Natarajan, MD MMSc, 185 Cambridge Street, CPZN 3.184, Boston, MA 02114, @pnatarajanmd, Jacques Fellay, MD PhD, EPFL-SV-GHI, Station 19, 1015 Lausanne, @jacquesfellay. denotes equal contribution (Alexander G. Bick, Konstantin Popadin, Pradeep Natarajan, Jacques Fellay).

## Abstract

People living with human immunodeficiency virus (PLWH) have significantly increased risk for cardiovascular disease in part due to inflammation and immune dysregulation. Clonal hematopoiesis of indeterminate potential (CHIP), the age-related acquisition and expansion of hematopoietic stem cells due to leukemogenic driver mutations, increases risk for both hematologic malignancy and coronary artery disease (CAD). Since increased inflammation is hypothesized to be both a cause and consequence of CHIP, we hypothesized that PLWH have a greater prevalence of CHIP. We searched for CHIP in multi-ethnic cases from the Swiss HIV Cohort Study (SHCS, n=600) and controls from the Atherosclerosis Risk in the Communities study (ARIC, n=8,111) from blood DNA-derived exome sequences. We observed that HIV is associated with increased CHIP prevalence, both in the whole study population and in a subset of 230 cases and 1002 matched controls selected by propensity matching to control for demographic imbalances (SHCS 7%, ARIC 3%, p=0.005). Additionally, unlike in ARIC, *ASXL1* was the most commonly implicated mutated CHIP gene. We propose that CHIP may be one mechanism through which PLWH are at increased risk for CAD. Larger prospective studies should evaluate the hypothesis that CHIP contributes to the excess cardiovascular risk in PLWH.

## Introduction

As current treatments have rendered human immunodeficiency virus (HIV) a chronic condition, coronary artery disease has emerged as a major source of morbidity in people living with human immunodeficiency virus (PLWH). Inflammation and immune dysregulation likely accelerate CAD risk among PLWH.^1^ Recently, ‘clonal hematopoiesis of indeterminate potential’ (CHIP), the age-related acquisition and expansion of leukemogenic mutations (primarily in *DNMT3A, TET2, ASXL1, JAK2*) in white blood cells, was found to increase risk for both hematologic malignancy^2,3^ and CAD^4,5^ among asymptomatic individuals in the general population. The proatherogenic mechanisms for CHIP included heightened inflammation.^4,6^ Given converging mechanisms promoting CAD risk and increased hematologic malignancy risk among PLWH, we tested the hypothesis that HIV-infected individuals have heightened prevalence of CHIP.

## Methods

We identified CHIP in a multi-ethnic sample of 600 PLWH who had available exome sequences from the Swiss HIV Cohort Study (SHCS), aged 21-83. The SHCS is a multicenter, prospective observational study for interdisciplinary HIV research^7^. Established in 1988, the SHCS currently comprises more than 20,000 PLWH with median 51 years of age. Samples of 600 patients, used for exome sequencing, were chosen randomly in terms of gender, age, category of transmission, as well as HIV management and control.^8^

We utilized a set of 8111 individuals with available exome sequences from the Atherosclerotic Risk in the Community study (ARIC), aged 45-84 years, as population controls.^9^ The ARIC study is a prospective longitudinal investigation of the development of atherosclerosis and its clinical sequelae which enrolled 15,792 individuals aged 45 to 64 years at baseline.^10^ At study enrollment (1987-1989), the participants were selected by probability sampling from four United States communities: Forsyth County, North Carolina; Jackson, Mississippi; the northwestern suburbs of Minneapolis, Minnesota; and Washington County, Maryland.

CHIP was called in both exome sequenced cohorts using a previously described pipeline.^4,11^ Briefly, short read sequence data was aligned to the hg19 reference genome using the BWA-mem algorithm and processed with the Genome Analysis Toolkit MuTect2 tool to detect somatic variants.^12^ Identification of individuals with CHIP, used a pre-specified list of variants in 74 genes known to be recurrent drivers of myeloid malignancies.

As CHIP prevalence depends strongly on age, we performed a 1:5 case/control propensity matching on age, sex and self-reported ethnicity using nearest neighbor matching^13^ and requiring an exact match on age as implemented by the MatchIt package version 3.0.2 in R. Univariate Fisher’s exact test and multivariate logistic regression tested the association between HIV status and CHIP prevalence. Multivariate models were adjusted for age, sex, self-reported ethnicity, and smoking status. Analyses were performed in R version 3.6. A threshold of p<0.05 was considered statistically significant.

Written informed consent was obtained from all human participants by each of the studies with approval of study protocols by ethics committees at participating institutions. Secondary analysis of the data in this manuscript was approved by the Mass General Brigham Institutional Review Board. All relevant ethics committees approved this study and this work is compliant with all relevant ethical regulations.

## Results

We began by considering the fraction of CHIP across the entire SHCS PLWH cohort (N=600) and ARIC cohort (N=8111) (Figure 1). SHCS PLWH and ARIC participants had mean (SD) age 44 (11) and 57 (6) years (p=1.8 x 10^-167^), were 25% and 56% female (p=1.9 x 10^-46^), and were 95% and 74% of European ancestry (p=5.2 x 10^-36^) respectively. With adjustment for age, age^2^, sex and ethnicity, we observed a significant association between HIV case status and CHIP (OR: 1.77, 95% CI: 1.33-2.21, p=0.02).

**Figure 1:**
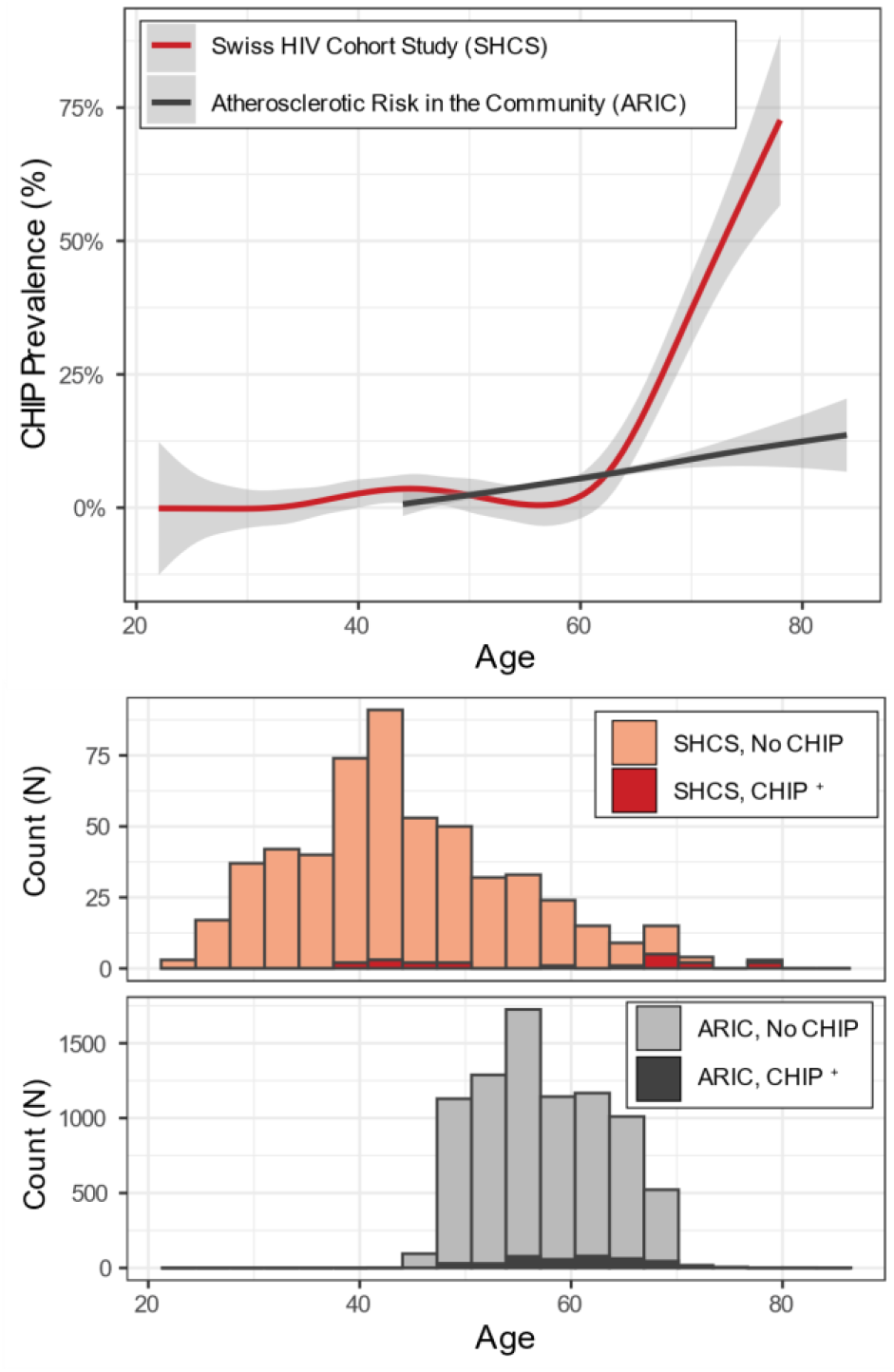
CHIP prevalence in Swiss HIV Cohort Study and Atherosclerotic Risk in the Community Study. Upper panel: fraction of cohort observed to have CHIP over time fit with a general additive model spline. 95% confidence interval displayed as shaded area. Lower panel: Count of number of individuals with and without CHIP binned by age of time of blood sampling across entire sequenced cohort.

Give the overall demographic imbalances, we pursued a propensity matching strategy and matched datasets by age, gender and ethnicity. Propensity matching analyses yielded a set of 230 PLWH cases and 1002 ARIC population controls. Neither age nor sex, differed significantly between the matched cohorts (Table 1) and the standardized mean difference across age, sex and self-reported ethnicity were all less than 0.1 indicative of adequate matching. In this subset, CHIP was detected in 7% of exomes from PLWH, but only 3% of the controls (Table 1, univariate p=0.005; multivariate p=0.004). Of note, the statistical association strengthened despite a significantly decreased sample size, likely due to the exclusion of younger SHCS PLWH, who are less likely to have CHIP. Depth of coverage of the four most common CHIP genes (*DNMT3A, TET2, ASXL1, JAK2*), when incorporated into the multivariate logistic regression model, did not affect the results.

**Table 1:**
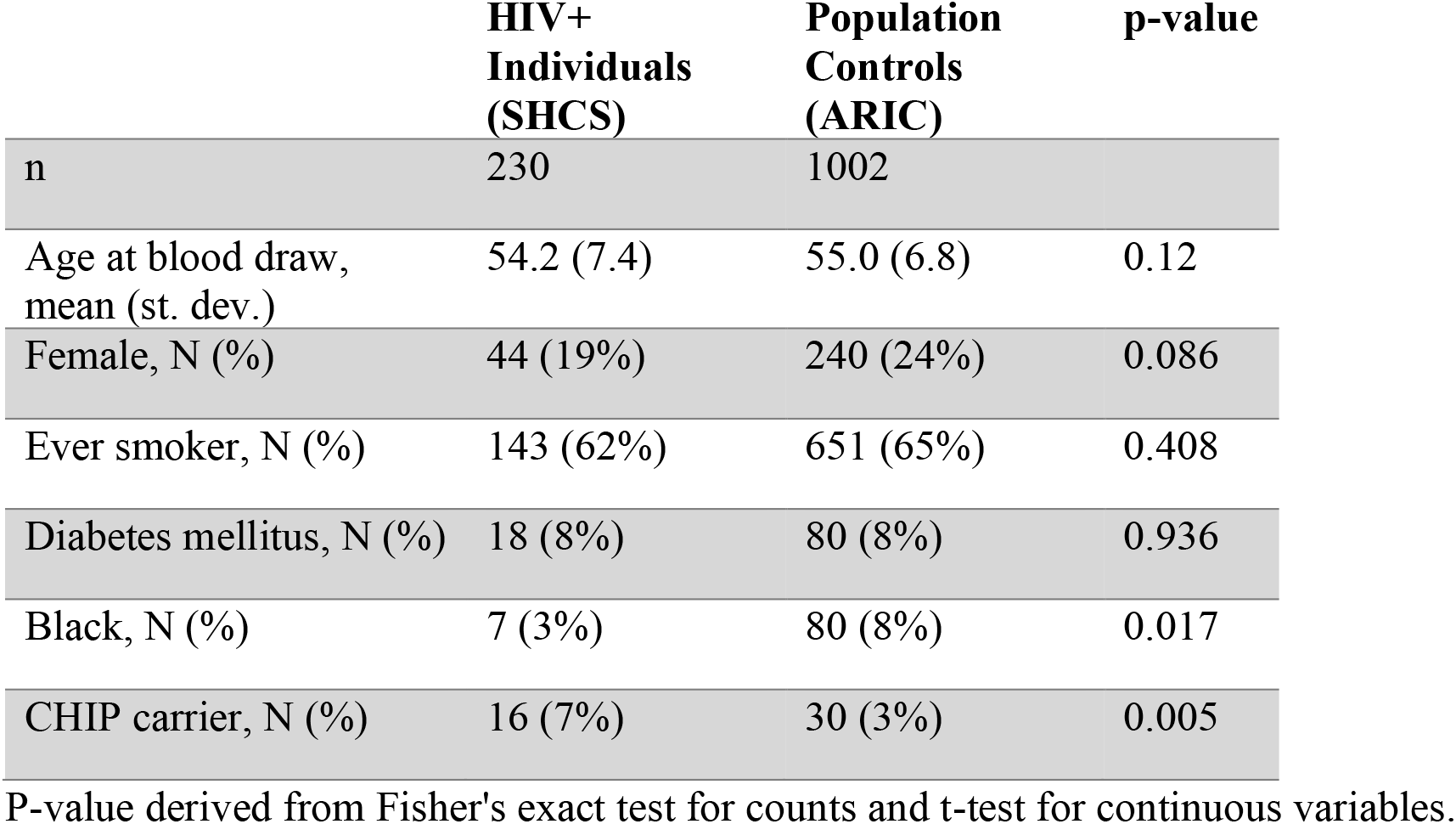
Demographics and CHIP association in matched samples.

|  | **HIV+**<br>**Individuals**<br>**(SHCS)** | **Population**<br>**Controls**<br>**(ARIC)** | **p-value** |
| --- | --- | --- | --- |
| n | 230 | 1002 |  |
| Age at blood draw,<br>mean (st. dev.) | 54.2 (7.4) | 55.0 (6.8) | 0.12 |
| Female, N (%) | 44 (19%) | 240 (24%) | 0.086 |
| Ever smoker, N (%) | 143 (62%) | 651 (65%) | 0.408 |
| Diabetes mellitus, N (%) | 18 (8%) | 80 (8%) | 0.936 |
| Black, N (%) | 7 (3%) | 80 (8%) | 0.017 |
| CHIP carrier, N (%) | 16 (7%) | 30 (3%) | 0.005 |

P-value derived from Fisher's exact test for counts and t-test for continuous variables.

The limited sample size precluded inference on the association of HIV status with specific CHIP driver genes, however we observed differences in the genes most likely to carry CHIP mutations between PLWH and population controls. The most common CHIP gene in the SHCS was *ASXL1* (13 out of 27 CHIP mutations, 48%) followed by *TET2* (8 out of 27 CHIP mutations, 30%) and *DNMT3A* (5 out of 27 CHIP mutations, 19%). Overall this distribution was inverted from the control cohort where CHIP mutations were more frequent in *DNMT3A*, followed by *TET2* and *ASXL1*. In total, 22 PLWH had a single CHIP mutation, while one individual had 2 mutations and one individual had 3 mutations.

Within the full PLWH cohort (N=600) we considered additional phenotypes, which might be a cause or consequence of CHIP. First, we observed a trend toward an increase in CAD among CHIP carriers (Fisher’s exact test OR: 2.99, p = 0.068). Second, we observed that duration of antiretroviral therapy (ART) was twice as long in CHIP carriers versus non-carriers (ART mean (st. dev.) ART 2675 [1850] days vs 1322 [1454] days in carriers vs non-carriers respectively; p = 0.0004, Mann-Whitney U test). This association was directionally concordant after adjusting for patient age in multiple logistic regression (p = 0.066) or and remained significant after matching of 24 CHIP carriers with 24 non-carriers by age (p = 0.042 paired Mann-Whitney U test). It is important to note that although ART duration positively correlates with the total duration of HIV infection (Spearman’s rho = 0.58, p=2.0 x 10^-54^,N = 600), the total duration of HIV infection is not associated with CHIP p = 0.452; paired Mann-Whitnmey U test on matched CHIP carriers and non-carriers, p = 0.22]

## Discussion

We here report that HIV associates with increased prevalence of CHIP, a recently recognized risk factor for blood cancer and CAD. In the present samples, we identify at least 2-fold enrichment of CHIP among PLWH versus controls when considering known factors predisposing to CHIP.

HIV infection is linked to accelerated biologic aging and chronic low-grade inflammation, providing a fertile substrate for CHIP development. Our study is consistent with another recent study that showed that HIV leads to a greater risk of myelodysplastic syndrome (MDS), a downstream consequence of CHIP and precursor to myeloid malignancy.^14^ Furthermore, similar to the gene distribution in MDS, we find a greater relative prevalence of *ASXL1* mutations among PLWH compared to controls. Of note, while cigarette smoking selects for *ASXL1* clonal hematopoiesis^15^, our cohort of PLWH still had an increased prevalence of *ASXL1* mutations compared to the control cohort despite being well balanced for smoking status across cohorts.

HIV infection may promote CHIP development through various mechanisms, including induced immunodeficiency, chronic immune activation from antigenic simulation, as well as increased prevalence of tobacco smoking and other co-morbid conditions. HIV may induce CHIP also through the ART which can either increase the rate of somatic mutagenesis or change the fitness landscape of hematopoietic stem cells or decrease effective population size of blood cells. HIV may also modify the selective coefficients of specific CHIP mutations. A recent model proposed that many of the CHIP mutations increase cell fitness, ensuring their proliferation with age.^16^ The relative contribution of these factors to CHIP risk, including in the context of spontaneous viral control and antiretroviral therapy, will require larger studies. An important limitation of the present study is its cross-sectional nature, but CHIP is highly unlikely to be a risk factor for HIV acquisition. The relative contribution of these factors to CHIP risk, including in the context of spontaneous viral control and antiretroviral therapy, will require larger studies. An important limitation of the present study is its cross-sectional nature, but CHIP is highly unlikely to be a risk factor for HIV acquisition.

We propose that CHIP may be one mechanism that elevates risk for CAD in PLWH. Further studies are required to evaluate the hypothesis that CHIP contributes to the excess cardiovascular risk associated with long-term HIV infection. CHIP may represent a unique opportunity for precision identification and targeting of CAD risk with particular relevance for HIV medicine.

## Data Availability

CHIP genetic variant callsets and associated participant level phenotype data used in this study are available to qualified investigators by application to the SHCS and ARIC.

## Acknowledgments

SHCS data are gathered by the Five Swiss University Hospitals, two Cantonal Hospitals, 15 affiliated hospitals and 36 private physicians (listed in http://www.shcs.ch/180-health-care-providers). The authors acknowledge the effort and commitment of SHCS participants, investigators, study nurses, laboratory personnel, and administrative assistance by the SHCS coordination and data center.

## Financial support

This study has been financed within the framework of the Swiss HIV Cohort Study, supported by the Swiss National Science Foundation (grant #177499), by SHCS project #860 and by the SHCS research foundation. This work was also supported by the Swiss National Science Foundation grant #175603 to JF. AGB is supported by a Burroughs Wellcome Foundation career award for medical scientists and a grant from the National Institute of Health Common Fund (DP5 OD029586). SG is supported by P30 DK040561 and U01HL123336. PN is supported by a Hassenfeld Scholar Award from the Massachusetts General Hospital, and grants from the National Heart, Lung, and Blood Institute (R01HL1427, R01HL148565, R01HL148050, and R01HL151283) and Fondation Leducq (TNE-18CVD04). Dr. Libby receives funding support from the National Heart, Lung, and Blood Institute (1R01HL134892), the American Heart Association (18CSA34080399), the RRM Charitable Fund, and the Simard Fund.

The Atherosclerosis Risk in Communities study has been funded in whole or in part with Federal funds from the National Heart, Lung, and Blood Institute, National Institutes of Health, Department of Health and Human Services (contract numbers HHSN268201700001I, HHSN268201700002I, HHSN268201700003I, HHSN268201700004I and HHSN268201700005I). The authors thank the staff and participants of the ARIC study for their important contributions. Funding support for “Building on GWAS for NHLBI-diseases: the U.S. CHARGE consortium” was provided by the NIH through the American Recovery and Reinvestment Act of 2009 (ARRA) (5RC2HL102419). Sequencing was carried out at the Baylor College of Medicine Human Genome Sequencing Center (U54 HG003273 and R01HL086694).

## Competing Interests

Dr. Libby is an unpaid consultant to, or involved in clinical trials for Amgen, AstraZeneca, Baim Institute, Beren Therapeutics, Esperion, Therapeutics, Genentech, Kancera, Kowa Pharmaceuticals, Medimmune, Merck, Norvo Nordisk, Merck, Novartis, Pfizer, Sanofi-Regeneron. Dr. Libby is a member of scientific advisory board for Amgen, Corvidia Therapeutics, DalCor Pharmaceuticals, Kowa Pharmaceuticals, Olatec Therapeutics, Medimmune, Novartis, and XBiotech, Inc. Dr. Libby’s laboratory has received research funding in the last 2 years from Novartis. Dr. Libby is on the Board of Directors of XBiotech, Inc. Dr. Libby has a financial interest in Xbiotech, a company developing therapeutic human antibodies. Dr. Libby’s interests were reviewed and are managed by Brigham and Women’s Hospital and Partners HealthCare in accordance with their conflict of interest policies. Dr Natarajan reported grants from Amgen during the conduct of the study and grants from Boston Scientific; grants and personal fees from Apple; personal fees from Novartis and Blackstone Life Sciences; and other support from Vertex outside the submitted work.

## Members of the Swiss HIV Cohort Study

Aebi-Popp K, Anagnostopoulos A, Battegay M, Bernasconi E, Böni J, Braun DL, Bucher HC, Calmy A, Cavassini M, Ciuffi A, Dollenmaier G, Egger M, Elzi L, Fehr J, Fellay J, Furrer H, Fux CA, Günthard HF (President of the SHCS), Haerry D (deputy of “Positive Council”), Hasse B, Hirsch HH, Hoffmann M, Hösli I, Huber M, Kahlert CR (Chairman of the Mother & Child Substudy), Kaiser L, Keiser O, Klimkait T, Kouyos RD, Kovari H, Ledergerber B, Martinetti G, Martinez de Tejada B, Marzolini C, Metzner KJ, Müller N, Nicca D, Paioni P, Pantaleo G, Perreau M, Rauch A (Chairman of the Scientific Board), Rudin C, Scherrer AU (Head of Data Centre), Schmid P, Speck R, Stöckle M (Chairman of the Clinical and Laboratory Committee), Tarr P, Trkola A, Vernazza P, Wandeler G, Weber R, Yerly S.

